# Impact of remote-monitored home non-invasive ventilation on patient outcomes: a retrospective cohort study

**DOI:** 10.1101/2024.04.11.24305702

**Authors:** Charlotte Levey, Maureen Manthe, Anna Taylor, Maryam Sahibqran, Eve Walker, Grace McDowell, Eric Livingston, Adam V. Benjafield, Chris Carlin

## Abstract

**Introduction:** Use of home non-invasive ventilation (NIV) to treat persistent hypercapnic respiratory failure in patients with stable chronic obstructive pulmonary disease (COPD) effectively reduces readmission rates and mortality compared with standard therapy. Traditional workflows around the initiation and management of NIV include elective admission for therapy initiation and frequent face-to-face clinic visits for follow-up, but use of telemedicine offers an alternative approach.

**Aim:** This retrospective cohort study evaluated the clinical efficacy and health resource use impact of a remote monitoring approach to the initiation and monitoring of home NIV.

**Methods:** Individuals with COPD, COPD-obstructive sleep apnoea or obesity-related respiratory failure who were started on remote-monitored home NIV from July 2016 to December 2020 were included. Data were obtained from electronic health records. The primary outcome was healthcare utilisation (hospital admissions and average number of bed days) in the 12 months after versus 12 months before starting NIV; secondary endpoints included 2-year survival and time to readmission, and blood gas analysis.

**Results:** In the 12 months after versus before NIV initiation, there was a significant reduction in the mean number of admissions (1.0±2.1 vs. 1.4±2.1; p<0.0001) and occupied bed days (9.6±26.8 vs. 17.2±27.5; p<0.0001); results were consistent across NIV indications. Time to first readmission (hazard ratio [HR] 2.11, 95% confidence interval [CI] 1.58–2.8; p<0.001) and time to death (HR 2.25, 95% CI 1.51–3.34; p<0.0001) were significantly worse in NIV non-users versus users, but did not differ by deprivation quintile. Blood gas analysis showed that NIV significantly reduced carbon dioxide pressure and bicarbonate compared with before NIV.

**Conclusions:** A technology-assisted service model for the remote initiation and monitoring of home NIV therapy for individuals with chronic hypercapnic respiratory failure was feasible, had a beneficial effect on healthcare utilisation and outcomes, and offset typical adverse relative survival outcomes associated with deprivation.

**KEY MESSAGES:** *What is already known on this topic:* When given at adequate pressures that ensure sufficient reduction in carbon dioxide pressure, home non-invasive ventilation (NIV) is an effective and well tolerated treatment for chronic hypercapnic respiratory failure in individuals with chronic obstructive pulmonary disease or obesity-related respiratory failure.

*What this study adds:* This study showed the feasibility and effectiveness of a remote monitoring approach to the initiation and management of home NIV therapy in a real-world setting.

*How this study might affect research, practice or policy:* As well as improving outcomes in appropriately selected individuals, the initiation and management of home NIV therapy using remote monitoring has the potential to improve workflow, equitably enhance access to and outcomes from treatment, and provide a rich continuous dataset that could facilitate derivation of actionable artificial intelligence insights to support proactive care interventions.

## INTRODUCTION

Chronic obstructive pulmonary disease (COPD) is a common chronic condition that is associated with major public health and economic impact.^1^ ^2^ COPD also has a significant impact on affected individuals, having a similar symptom burden to that of cancer,^3^ and a comparable impact on health status to diabetes or cardiovascular disease.^4^ Approximately one-quarter of all individuals with stable COPD have persistent hypercapnia, which is associated with poor prognosis, including hospital readmission rates of 80% and 1-year mortality rates of 50% after an index acute hypercapnic respiratory failure episode.^5^

Home non-invasive ventilation (NIV) used at pressures that substantially reduce carbon dioxide levels improves outcomes in individuals with COPD and persistent hypercapnia.^6^ ^7^ Köhnlein and colleagues have shown that the addition of high-intensity NIV to standard therapy reduced 1-year mortality by 76% and significantly improved health-related quality of life compared with standard therapy alone in stable COPD patients with chronic hypercapnic respiratory failure.^7^ In the randomised, controlled Home Oxygen Therapy-Home Mechanical Ventilation (HOT-HMV) study, improved outcomes with home NIV plus oxygen therapy compared with oxygen therapy alone for patients with severe COPD and persistent hypercapnia after hospitalisation for an acute disease exacerbation included a 51% decrease in the risk of hospital readmission or death, a 74% reduction in the risk of hospital readmission in the first 28 days, and a 34% decrease in the exacerbation rate.^8^

Initiation of home NIV within an index acute episode is established standard of care for patients with chronic hypercapnic respiratory failure (PaCO_2_ >7kPa) related to obesity (obesity-related respiratory failure [ORRF]), where pooled outcomes indicate a 4-year mortality rate of 50% without therapy, which is normalised by optimised respiratory support.^9 10^. Home NIV is also recommended for patients with chronic hypercapnic respiratory failure in the context of COPD who have suspected or confirmed overlapping obstructive sleep apnoea (COPD-OSA).

Traditional workflows around the initiation and management of NIV have included elective admission for therapy initiation and frequent face-to-face clinic visits for follow-up. Increasing prevalence rates of both COPD and obesity is resulting in increasing demand on home NIV services, requiring adaptation of service models to improve patient experience, clinical efficiency and capacity.^11^ In our organisation (The National Health Service [NHS] Greater Glasgow & Clyde [GG&C]) we have progressively adopted the routine use of remote monitoring for the set-up and continuing management of home NIV therapy since 2016.

Cost-free access to unlimited duration remote-monitoring data for NIV and continuous positive airway pressure has been mandated for suppliers in the NHS in Scotland since 2018, allowing us to expand our home NIV service provision. A preliminary retrospective data analysis showed that this approach was feasible for individuals with COPD and was associated with longer time to readmission or death over 12 months’ follow-up.^12^ However, further exploration of the feasibility and efficacy of this remote-managed NIV service model in a larger cohort of patients with COPD and in patients with other chronic hypercapnic respiratory failure aetiologies is needed. Key considerations are whether this service model achieves acceptable rates of home NIV therapy continuation, effectively controls hypercapnic respiratory failure, improves outcomes in line with previous clinical trials, and offsets typical access challenges and adverse outcomes associated with deprivation.

This single-centre retrospective cohort study evaluated the clinical efficacy and health resource use impact of a remote monitoring approach to the initiation and monitoring of home NIV in individuals with COPD, COPD-OSA or ORRF.

## METHODS

### Study design and participants

This retrospective cohort study was conducted within NHS GG&C. Individuals who were started on remote-monitored home NIV over the period July 2016 to December 2020 (identified from the AirView™ NIV database) were eligible; those who started on, or were transitioned to, continuous positive airway pressure (CPAP) therapy were excluded.

Individuals with neuromuscular disease, chest wall disease, and central nervous system diseases were also excluded because they are managed via a different pathway. Eligibility for home NIV therapy was determined based on established guidelines.^13^ ^14^ Data access, handling and aggregation of de-identified data for this service evaluation and publication was approved by NHS GG&C Caldicott Guardian.

### Treatment

Home NIV for chronic hypercapnic respiratory failure was initiated and followed up based on the NHS GG&C clinical protocol (Figure 1). Home NIV was provided using a Lumis 150 ST-A device (ResMed) in intelligent volume-assured pressure support (iVAPS) auto-expiratory positive airway pressure (auto-EPAP) mode or spontaneous timed (ST) mode with patient-consented remote monitoring via the AirView™ platform (ResMed).

**Figure 1.**
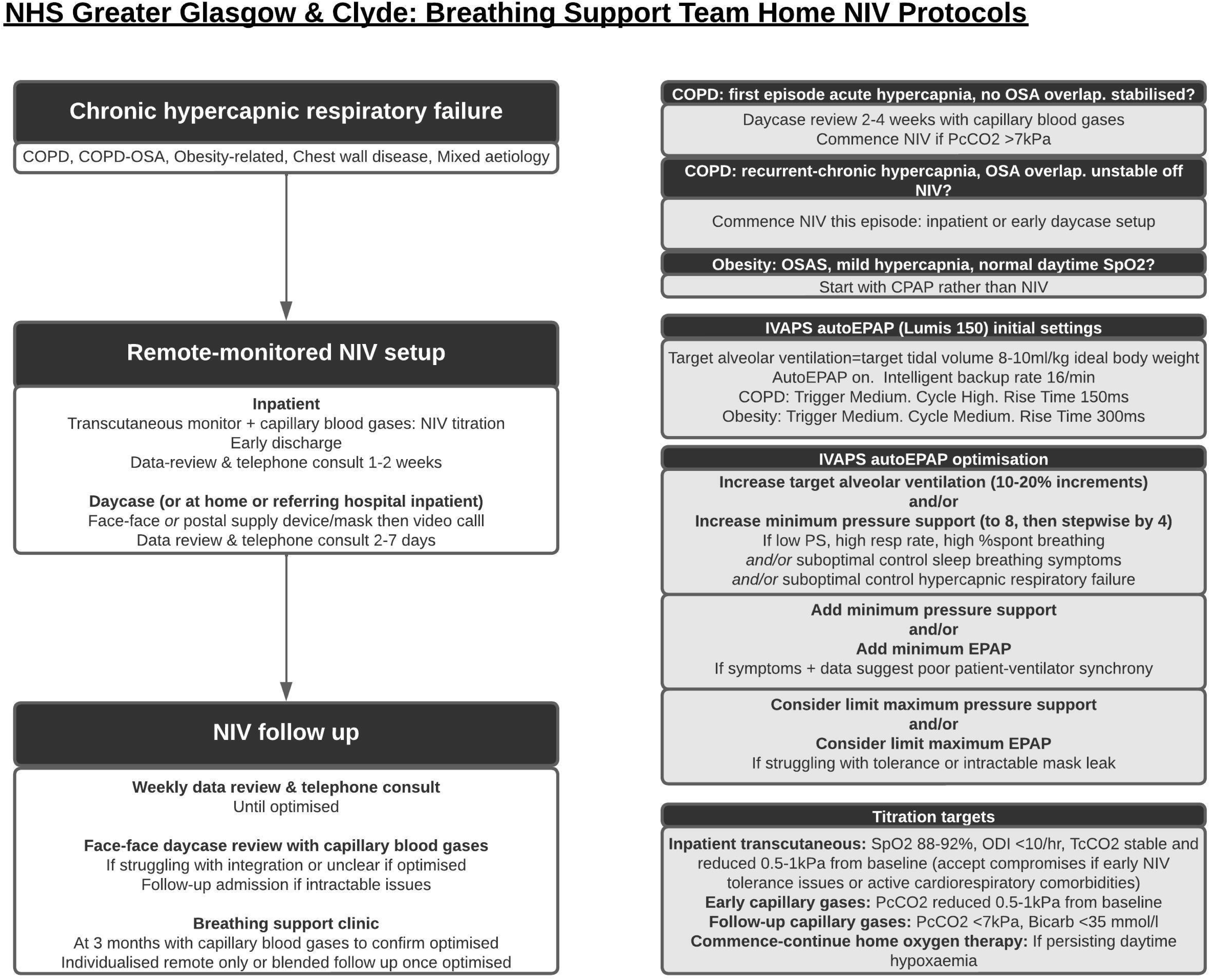
Protocol for the initiation and follow-up of home non-invasive ventilation. Bicarb, bicarbonate; COPD, chronic obstructive pulmonary disease; EPAP, expiratory positive airway pressure; IVAPS, intelligent volume-assured pressure support; NIV, non-invasive ventilation; ODI, oxygen desaturation index; OSA, obstructive sleep apnoea; PcCO2, capillary pressure of carbon dioxide; PS, pressure support; SpO2, oxygen saturation; TcCO2, transcutaneous pressure of carbon dioxide.

### Definitions

COPD was defined using the Global Initiative for Chronic Obstructive Lung Disease (GOLD) criteria.^14^ Individuals with ORRF had a body mass index (BMI) of >30 kg/m^2^ with chronic hypercapnic respiratory failure and/or nocturnal hypoventilation and/or recurrent episodes of acute hypercapnic respiratory failure with symptomatic or polygraphy-confirmed sleep-disordered breathing (as per the National Institute for Health and Care Excellence definition^15^). Individuals with COPD-OSA had COPD (GOLD criteria^14^) plus polygraphy-confirmed OSA or BMI >30 kg/m^2^ with symptoms consistent with OSA syndrome and a daytime PaCO_2_ of >7kPa.

### Assessments and follow-up

Electronic health records (EHRs) were reviewed to obtain patient demographic data, primary diagnosis and indication for NIV, the date, location and nature of NIV set-up, whether NIV use was continued after set-up, the number of respiratory-related hospital admissions in the year before and the year after NIV initiation, date of readmission, date of death, and blood gas analysis results at baseline and follow-up (where available in the EHR). Individualised follow-up was performed as per clinical routine, meaning that individuals with satisfactory progress and remote monitoring data would not have follow-up blood gases. In addition, the impact of the COVID-19 pandemic meant that the number of face-to-face follow-up visits was lower than normal. Furthermore, the majority of individuals who discontinued NIV did not attend face-to-face follow-up (and therefore also did not have blood gases determined).

Data for follow-up analyses were censored on 22 February 2023. NIV continuation or discontinuation at 12 months was determined from clinical notes. AirView™ usage data were not available for this retrospective evaluation but would typically have been considered as part of follow-up and reflected in the decision to continue or discontinue NIV. The number and nature of follow-up reviews, changes in NIV settings, and the NIV interface are not systematically recorded in the EHR and were therefore not available for analysis.

### Outcomes

The primary outcome was healthcare utilisation based on the number of hospital admissions and the average number of bed days in the 12 months after starting NIV versus the 12 months before NIV initiation; these endpoints were also related to the indication for NIV, and ongoing usage of NIV at 12 months after therapy initiation. Secondary endpoints included 2-year survival and time to readmission, both related to observed NIV usage and Scottish Index of Multiple Deprivation (SIMD),^16^ and blood gas analysis data (PaCO_2_ and bicarbonate) at baseline and clinic follow-up.

### Statistical analyses

Baseline data are presented as mean, median, count, percentage, and/or range with standard deviation or interquartile range, as appropriate. The mean frequency of hospital admissions and average occupied bed days in the 12 months before and after NIV initiation were analysed using violin boxplots (for the overall population and in subgroups based on the indication for NIV) and values were compared using the Wilcoxon signed rank test. Survival outcome and time to readmission were evaluated using Kaplan-Meier survival analysis and compared with log-rank tests (for the overall cohort, by NIV usage, and SIMD quintile). Blood gas measurements were analysed at baseline and clinic follow-up using slope graphs and the Wilcoxon test. All statistical analyses were conducted using R (version 4.2.2) and GraphPad Prism v9 (GraphPad Software, San Diego, USA).

## RESULTS

### Study population

An overview of the remote-monitored home NIV cohort is shown in Figure 2. A total of 362 individuals were included in the analysis (274 users of NIV and 88 non-users of NIV) (Table 1). The most common indication for home NIV was ORRF, and 46% of the study population had home NIV initiated during an acute inpatient episode (Figure 2). Overall, 88 individuals started on NIV discontinued therapy during the first year (24%); this proportion was higher in the subgroup who had NIV setup during an acute inpatient episode (28%) than in the subgroup who had NIV setup on an elective home basis (21%). A high proportion of patients (45%) initiated on remote-monitored NIV were resident in SIMD 1 deprived areas (Figure S1).

**Figure 2.**
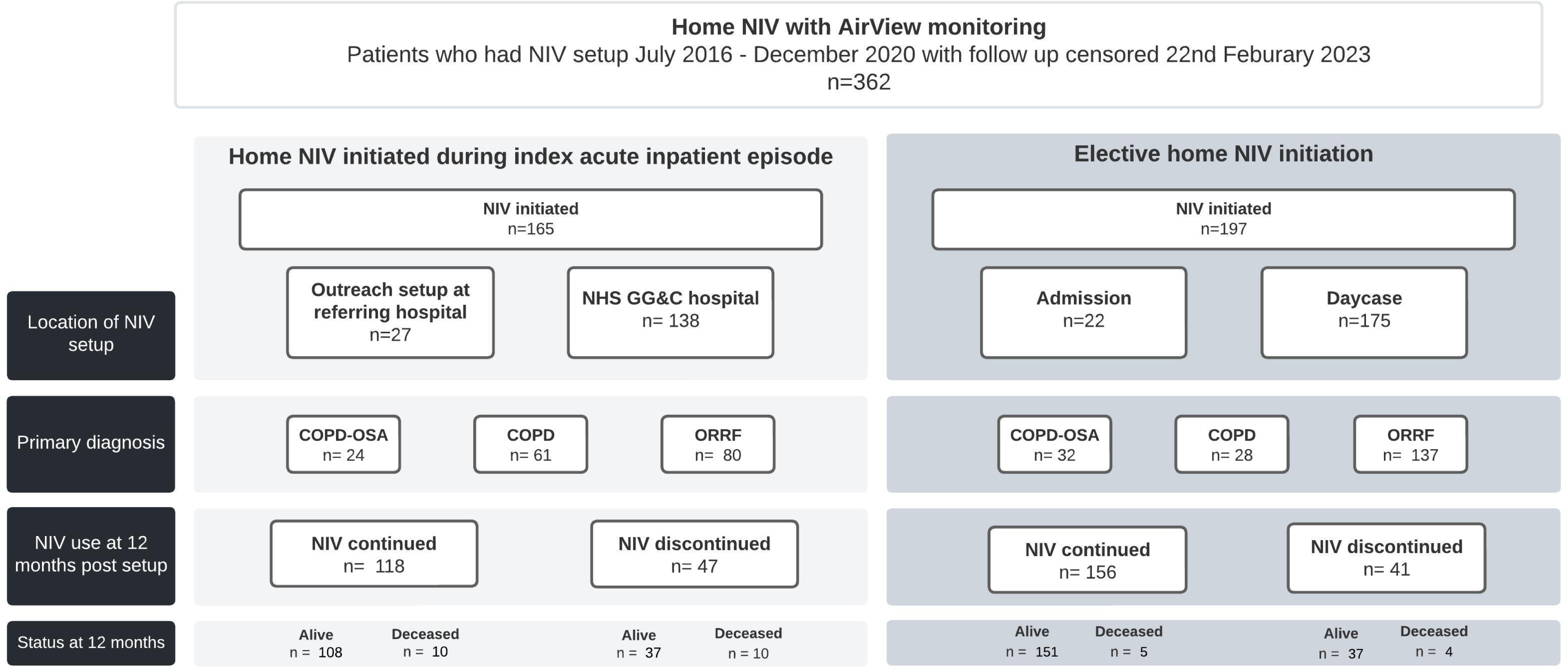
Overview of National Health Service Greater Glasgow & Clyde (NHS GG&C) remote-monitored home NIV cohort. COPD, chronic obstructive pulmonary disease; NIV, non-invasive ventilation; ORRF, obesity-related respiratory failure; OSA, obstructive sleep apnoea.

**Table 1.** Baseline characteristics of the remote-monitored home NIV cohort.

|  | **Total**<br>**(n=362)** | **NIV users**<br>**(n=274)** | **NIV non-users**<br>**(n=88)** |
| --- | --- | --- | --- |
| Male sex, n (%) | 155 (42.8) | 127 (46.4) | 28 (31.8) |
| Median age, years (range) | 62 (20–90) | 62 (20–90) | 62 (29–84) |
| Mean ± SD number of respiratory-related admissions in year prior to NIV setup | 1.4±2.1 | 1.2±1.8 | 2.4±2.7 |
| Mean pCO<sub>2</sub>, kPa (range) | 8 (3.5–16.8) | 8 (3.5–16.8) | 8.1 (5.2–15.1) |
| Mean bicarbonate, mmol/L (range) | 36 (22–61) | 36 (25–56) | 36 (22–61) |
| SIMD quintile, n (%) |  |  |  |
| 1 | 163 (45) | 123 (45) | 40 (45) |
| 2 | 92 (25) | 63 (23) | 29 (33) |
| 3 | 46 (13) | 39 (14) | 7 (8) |
| 4 | 40 (11) | 32 (12) | 8 (9) |
| 5 | 19 (5) | 16 (6) | 3 (3) |

NIV, non-invasive ventilation; pCO<sub>2</sub>, carbon dioxide pressure at initiation of home NIV (influenced in some patients by preceding acute NIV); SD, standard deviation; SIMD, Scottish Index of Multiple Deprivation where 1= most deprived quintile and 5 = least deprived quintile.

### Healthcare utilisation

Compared with the 12 months before NIV, there was a significant reduction in the mean number of admissions (1.0±2.1 vs. 1.4±2.1; p<0.0001) and the mean number of occupied bed days (9.6±26.8 vs. 17.2±27.5; p<0.0001) in the 12 months after NIV (Figure 3, Table S1).

**Figure 3.**
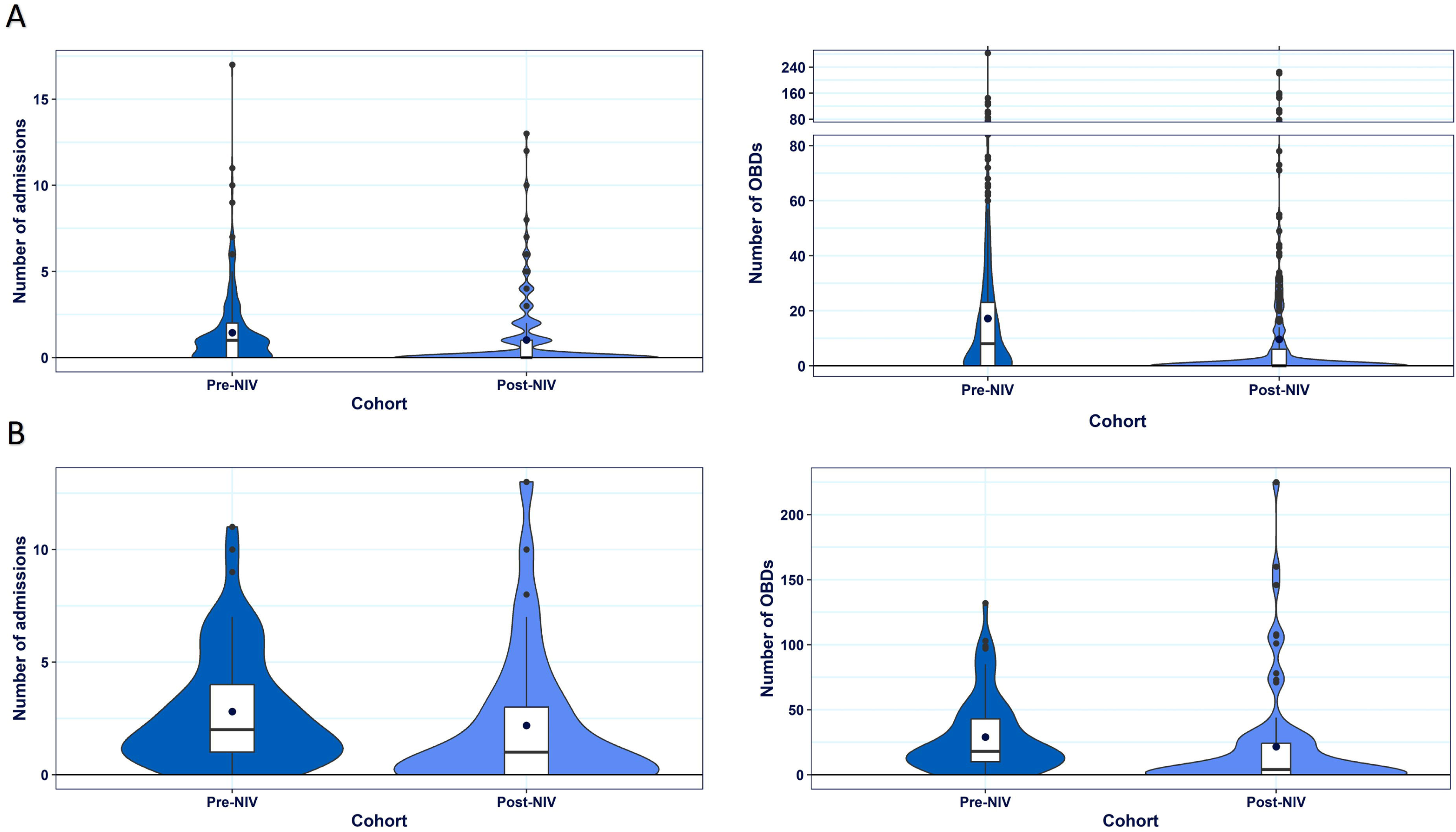
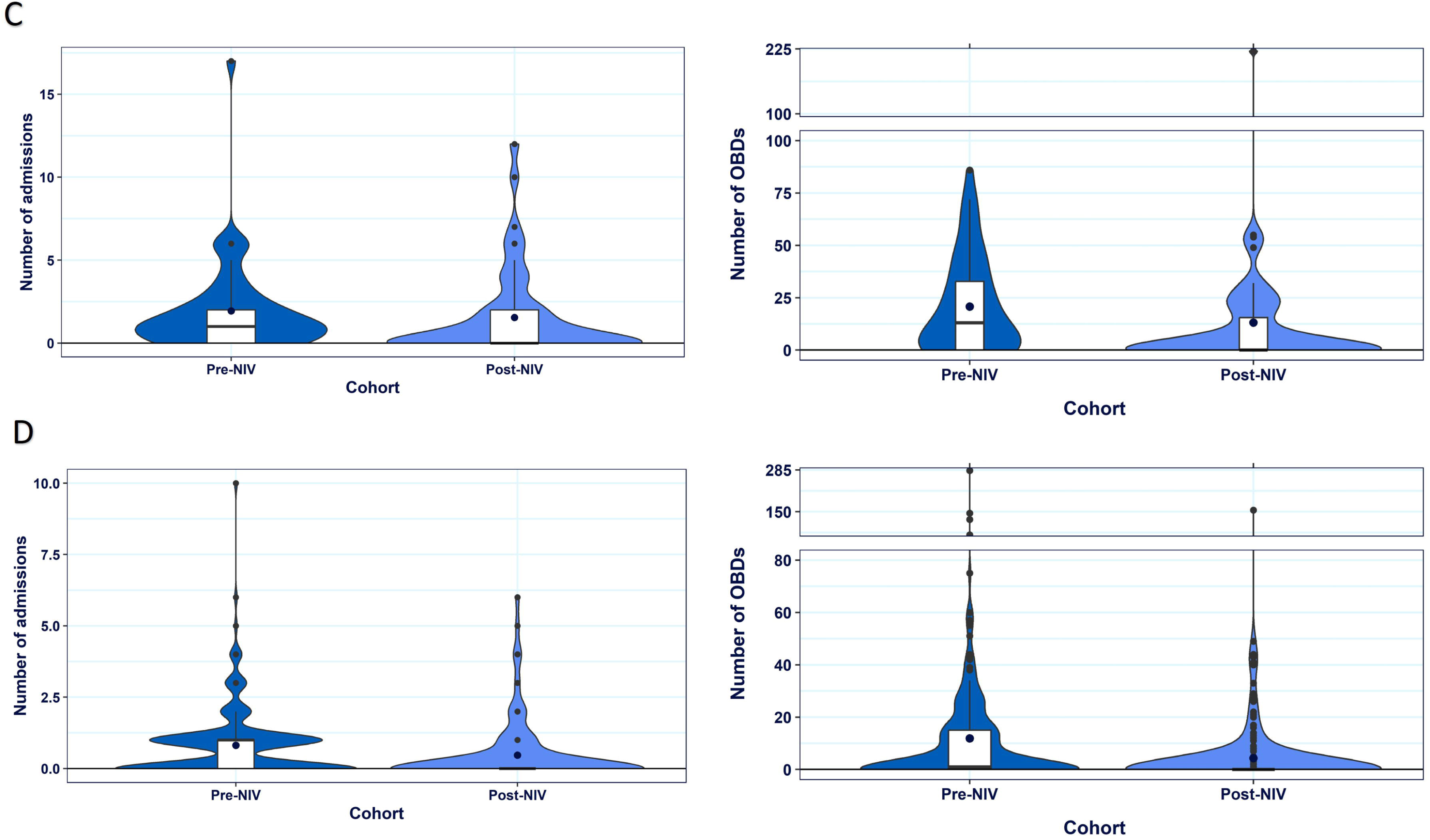
Violin box plots for the number of hospital admissions and number of occupied bed days in the 12 months before (pre) and after (post) initiation of non-invasive ventilation in (**A**) the total study population, (**B**) in patients with chronic obstructive pulmonary disease (COPD), (**C**) in patients with COPD-obstructive sleep apnoea and (**D**) in patients with obesity-related respiratory failure.

These significant reductions were seen across all indications for NIV, with the exception of the number of admissions in individuals with COPD-OSA (Figure 3, Table S1). Both measures of healthcare resource use decreased significantly in NIV users and NIV non-users (Table S1).

### Time to readmission or death

Median ime to first readmission (hazard ratio [HR] 2.11, 95% confidence interval [CI] 1.58– 2.8; p<0.001) and median time to death (HR 2.25, 95% CI 1.51–3.34; p<0.0001) were significantly worse in NIV non-users versus users (Figure 4, Table S2). Compared with the overall cohort, time to readmission or death was significantly worse in those with COPD (HR 2.31, 95% CI 1.78–3.00; p<0.0001) or COPD-OSA (HR 1.43, 95% CI 1.02–2.00; p=0.04), and significantly better in those with ORRF (HR 0.64, 95% CI 0.50–1.81; p=0.0002). Median time to death was significantly worse in those with COPD (HR 2.1, 95% CI 1.48–2.99; p<0.0001) and significantly better in those with ORRF (HR 0.62, 95% CI 0.44–0.89; p=0.0096) (Figure 4, Table S2). Median time to first readmission or death and median time to death did not differ significantly by SIMD quintile in the whole cohort (Figure 4, Table S2). However, median time to readmission or death in the SIMD 1 cohorts was better in NIV users versus non-users (1388 vs. 206 days) (Figure S2, Table S3).

**Figure 4.**
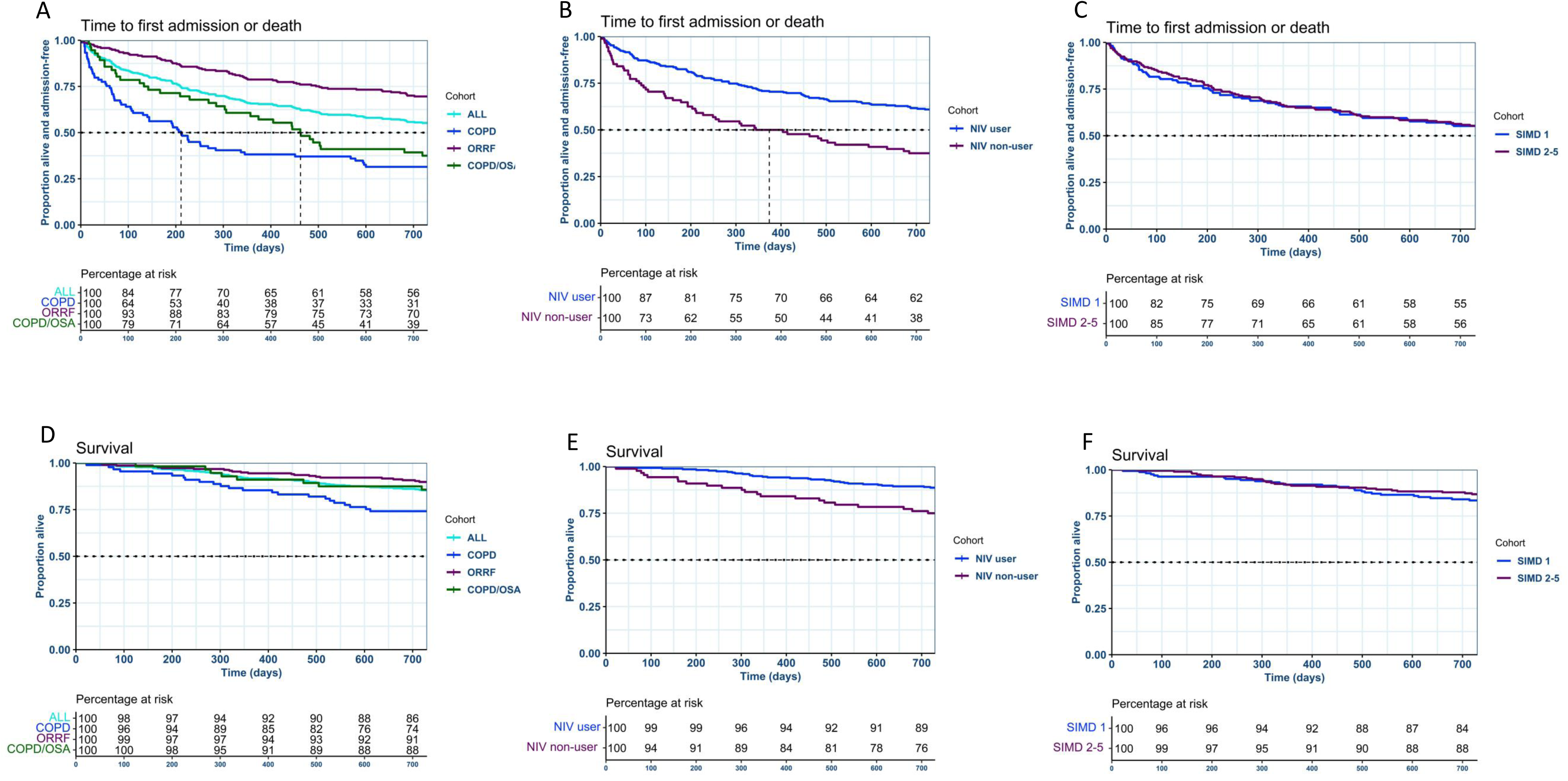
Kaplan-Meier survival plots with associated percentage at risk tables showing time to first readmission or death in subgroups based on the indication for NIV (**A**), use versus non-use of NIV (**B**) or Scottish Index of Multiple Deprivation quartile (**C**), and time to death in subgroups based on the indication for NIV (**D**), use versus non-use of NIV (**E**) or Scottish Index of Multiple Deprivation quartile (**F**). COPD, chronic obstructive pulmonary disease; NIV, non-invasive ventilation; ORRF, obesity-related respiratory failure; OSA, obstructive sleep apnoea; SIMD, Scottish Index of Multiple Deprivation.

### Blood gases

There was a significant reduction in both PaCO_2_ and bicarbonate during NIV therapy. The median change from baseline in PaCO_2_ was –1.7, –1.6 and −1.65 kPa, respectively, in participants with COPD, ORRF or COPD-OSA; corresponding median changes from baseline in bicarbonate in the three groups were –7, –6 and –6 mmol/L (Figure S3).

## DISCUSSION

This study showed the feasibility and effectiveness of a remote monitoring approach to the initiation and management of NIV therapy in a real-world setting. Additionally, it shows the feasibility and effectiveness in home NIV in individuals with COPD-OSA and ORRF. This contrasts with clinical studies, which have mainly reported results on individuals with hypercapnic COPD. In terms of healthcare resource use, there was a significant reduction in the number of admissions and the number of occupied bed days in the 12 months after initiation of NIV compared with the 12 months before NIV was started. Furthermore, NIV users had significantly longer time to readmission and time to death than NIV non-users.

In the current cohort, the median time to readmission or death was 946 days (about 31 months), which is substantially longer than the median time to readmission or death in the home NIV group of the HOT-HMV study (4.3 months).^8^ This may reflect differences in the study population, which covered three different indications in the current study (COPD, COPD-OSA and ORRF) compared with severe COPD only in the HOT-HMV study.

Individuals with COPD in the current study had lower survival rates than the overall population and the cohorts with COPD-OSA or ORRF. However, looking at individuals with COPD only, survival was better in our cohort compared with other similar cohorts.^8^ ^17^ Those with COPD-OSA had a shorter time to readmission than individuals with ORRF, but overall survival was similar in these two groups.

All individuals included in our analysis were either initiated on home NIV during an acute inpatient episode (46%), or at an elective day case outpatient visit (54%). Survival rates at 12 months were higher in the elective day case subcohort (95%) than in the acute inpatient set-up cohort (88%), possibly reflecting that the acute inpatient cohort was a sicker population.

The 1-year NIV discontinuation rate in our study was 24% (28% after inpatient setup, 21% after elective day case setup), similar to the 21% 1-year NIV discontinuation rate in a previous European analysis.^18^ Significant improvements in admissions and occupied bed days from the year before to the year after NIV were seen in NIV users and non-users at 12 months. However, numbers of both admissions and occupied bed days in the year before NIV were much higher in NIV nonusers versus users (2.3 vs. 1.2 admissions and 26.9 vs. 14.4 occupied bed days). The significant reductions seen in nonusers could represent regression to the mean, or be due to ancillary benefits and care-optimisation undertaken alongside the initiation of home NIV. The absolute mean number of both admissions (0.9±1.9 vs. 1.6±2.8) and occupied bed days (8.7±27.5 vs. 13.0±24.3) in the 12 months after NIV initiation was lower in NIV users than in non-users. Furthermore, there was a clear differentiation between users and non-users in terms of readmission and mortality outcomes. Non-users of NIV in our cohort had more than double the risk of readmission or death compared with users. This highlights the importance of continuing to strive to support long-term NIV usage, and to identify alternative interventions that improve outcomes in patients with COPD and chronic hypercapnic respiratory failure who are unable to continue home NIV despite support.

A high proportion of people in the current cohort resided in the most deprived areas (SIMD 1). This is consistent with reported associations between the prevalence of COPD and obesity and low socioeconomic status.^19–21^ This also mirrors the burden of disease within NHS GG&C (Figure S1) and Scotland. Time to readmission and death in the subgroup of individuals resident within SIMD 1 did not differ significantly from those resident in SIMD 2–5. This contrasts with the elevated standardised annual COPD mortality rates in SIMD 1 in the general population.^22^ There was, however, a difference in time to clinical events in SIMD 1 NIV non-users compared with NIV users, which was shorter in NIV non-users (Figure S2, Table S3). This is compatible with a direct benefit from continued home NIV use in these individuals. These findings suggest that an agile remote management-based home NIV service model could offset the typical therapy access and outcome inequalities that are associated with deprivation in patients with COPD and chronic hypercapnic respiratory failure.

Overall, the beneficial effects of remote-monitored home NIV on healthcare resource use in the current study add to the body of knowledge regarding the utility of telemedicine solutions for the initiation and management of home NIV. A previous randomised controlled trial conducted in The Netherlands used telemedicine to facilitate the initiation of chronic home NIV in individuals with stable COPD and chronic hypercapnia. In that trial, use of telemedicine was non-inferior to in-hospital NIV initiation with respect to reductions in PaCO_2_ and improvements in health-related quality of life. In addition, home initiation of NIV using telemedicine reduced costs by more than 50% compared with in-hospital NIV initiation.^23^

An economic analysis based on data from the HOT-HMV study, which used traditional methods for in-hospital NIV initiation, showed that the addition of home NIV to oxygen therapy had a 62% probability of being cost effective at a willingness-to-pay threshold of £30,000.^24^ Furthermore, another analysis using HOT-HMV study data showed that home NIV was cost-effective from a health system perspective. The cost per quality-adjusted life-year was £10,259, which is below the UK cost-effectiveness threshold and compares favourably with other common interventions used in COPD.^25^ Over the period covered by the current study, we were able to deliver a significant increase in NIV provision in Glasgow without any additional staffing. This is consistent with preliminary data from our centre showing no increase in nurse specialist team workload for remote NIV initiation.^12^ Given the cost savings and lower healthcare resource use that have been documented with remote-monitored home NIV initiation in the current study and a previous randomised trial,^23^ it is possible that this approach to the initiation and monitoring of home NIV might be more likely to be cost effective than the standard approaches that have previously undergone economic evaluation. The improved clinical efficiency, reduced patient travel and ability to interact proactively enabled by our service model are likely to add additional cost and sustainability benefits. These considerations require prospective evaluation in future studies.

Although providing useful information about the feasibility and impact of a remote monitoring approach to NIV therapy set-up and monitoring, several limitations need to be considered when interpreting our findings. There was a limited amount of data available in the EHRs used for this analysis. The means that we did not have information on factors such as smoking status, lung function, comorbidities, and living situation. Therefore, we were unable to understand or account for any associated biases. Participants in the current analysis were grouped by indication for NIV, but there are other factors that could influence outcomes, as shown by a recent study that found significant heterogeneity in health trajectories prior to the initiation of home NIV in France.^26^ Finally, it is important to note that the current study was conducted in a healthcare setting (NHS Scotland) where cost-free access to unlimited duration remote monitoring of CPAP and NIV data has been mandated since 2018. While this provided a large body of continuous long-term remote monitoring data to support clinical decision making and service delivery, the study findings are only applicable to areas with similar healthcare systems and need to be replicated in regions that might have different healthcare and reimbursement models. Additionally, because compliance and therapy usage data were not available for this retrospective cohort analysis, it is possible that some individuals in the NIV user group were only using the device infrequently at 12 months and that some individuals defined as having discontinued NIV subsequently resumed therapy.

In conclusion, this analysis showed that a technology-assisted model for the remote initiation and monitoring of home NIV therapy in individuals with chronic hypercapnic respiratory failure was feasible and effective. In this population, NIV therapy controlled hypercapnic respiratory failure, reduced healthcare resource use (admissions and hospital stay days), increased time to readmission or death, and offset the typical therapy access challenges and adverse relative survival outcomes that are seen in individuals resident in deprived areas. Additional research is needed to validate the current findings in different populations, but the data suggest that remote monitoring technology can play an important role in facilitating home NIV set-up and in therapy monitoring. Additionally, these types of service transformations provide the necessary service and digital infrastructure to explore the additional value that could be obtained from continuous NIV therapy data, including generating artificial intelligence-based predictive insights with early recognition of deteriorations, to allow proactive preventative interventions.

## Acknowledgements

Medical writing assistance for preparation of the manuscript was provided by Nicola Ryan, independent medical writer, funded by ResMed. We gratefully acknowledge the comprehensive contribution of the respiratory physiologist and nurse specialist teams in NHS Greater Glasgow and Clyde to the positive outcomes reported in this paper. They have enthusiastically adapted service models to realise benefits from assistive technologies, and their commitment to improving patient outcomes and providing realistic medicine is inspiring.

## Contributors

CL, MM, AT, MS, EW and CC analysed the data. All authors drafted and revised the manuscript. CC acts as guarantor and had full access to all of the data in the study and takes responsibility for the integrity of the data and the accuracy of the data analysis.

## Patient and public involvement

Due to the retrospective nature of the analyses, patients and/or the public were not involved in the study design and study enrolment.

## Patient consent for publication

Not applicable.

## Ethics approval

Data access, handling and aggregation of de-identified data for this service evaluation and publication was approved by NHS Greater Glasgow and Clyde Caldicott Guardian.

## Data availability statement

Data are available from the senior author on reasonable request.

