## Supplement for "Impact of remote-monitored home non-invasive ventilation on patient outcomes: a retrospective cohort study"

**Figure S1.** Scottish Index of Multiple Deprivation (SIMD) distribution breakdown for the study cohort and the local COPD population.

(Local COPD population data from the 2018-19 Public Health Scotland Atlas of Variation respiratory dashboard for COPD patients with 1–3 admissions per year; SIMD 1 indicates the most deprived quintile and SIMD 5 indicates the least deprived quintile).

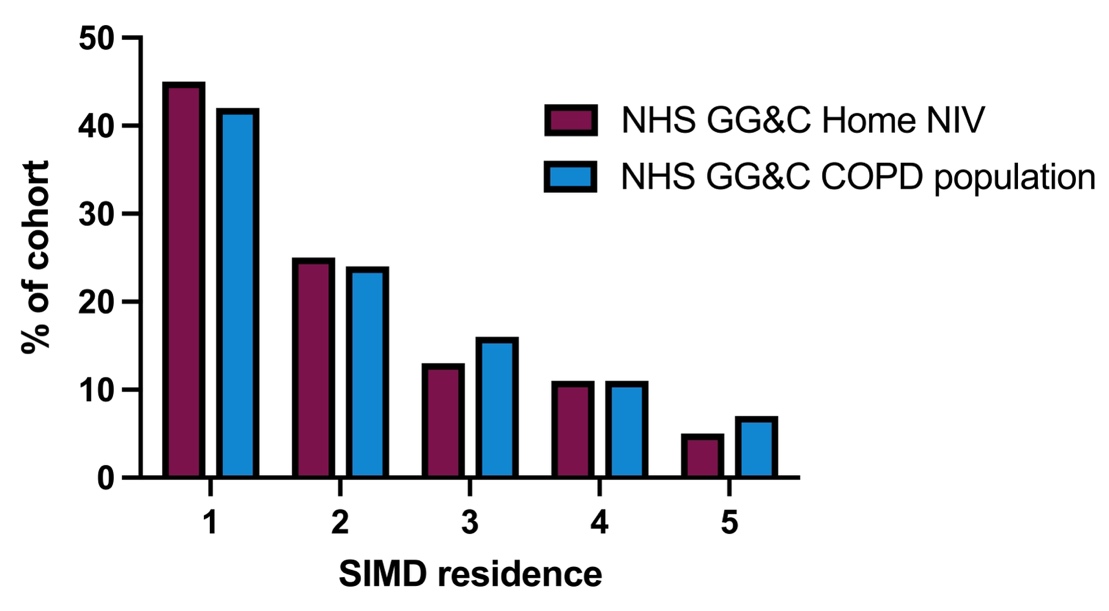

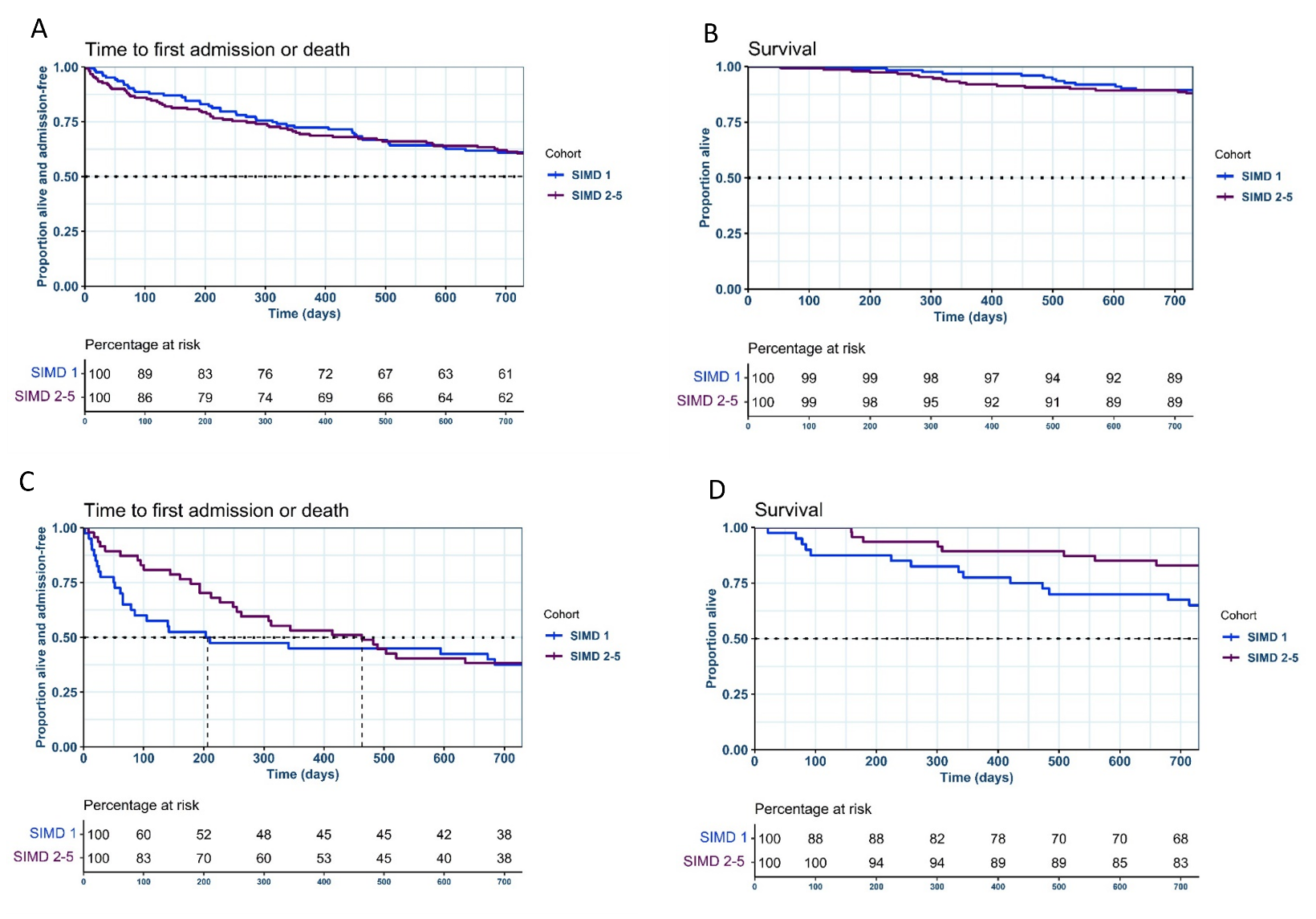
**Figure S2.** Kaplan-Meier survival plots with associated percentage at risk tables showing time to first readmission or death in subgroups based on Scottish Index of Multiple Deprivation (SIMD) quartile in NIV users at 12 months (A, B) and NIV non-uers at 12-months (C, D).

**Figure S3.** Individual changes in blood gas measurements from baseline to follow-up in users of home non-invasive ventilation (NIV) for participants who attended for face-to-face follow-up and had some or all blood gas analysis data inputted into their electronic health record by indication for NIV therapy; follow-up was performed ≥3 months after therapy initiation.

COPD, chronic obstructive pulmonary disease; OSA, obstructive sleep apnoea; PCO2, carbon dioxide pressure.

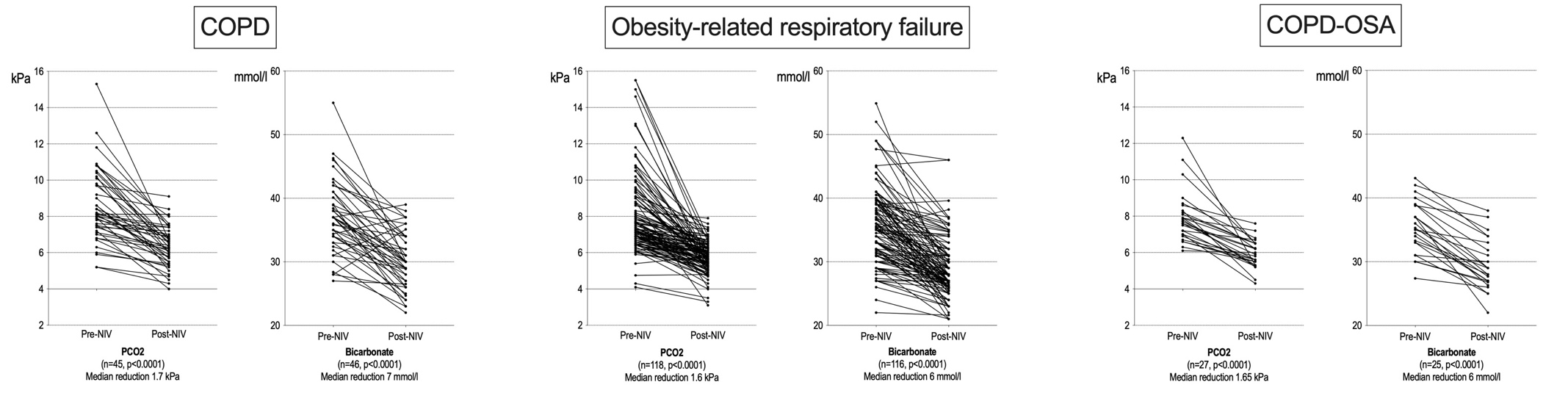

**Table S1.** Annual healthcare utilisation in the 12 months before and after initiation of home non-invasive ventilation.

|  | **Number alive at 12 months** | **Number of admissions** | | | **Occupied bed days** | | |
| --- | --- | --- | --- | --- | --- | --- | --- |
|  |  | **Before NIV** | **After NIV** | **p-value** | **Before NIV** | **After NIV** | **p-value** |
| All patients | 333 | 1.4 ±2.1 | 1.0±2.1 | <0.0001 | 17.2±27.5 | 9.6±26.8 | <0.0001 |
| COPD | 76 | 2.8±2.5 | 2.2±3.1 | 0.02 | 29.0±28.7 | 21.5±41.0 | 0.009 |
| ORRF | 205 | 0.8±1.2 | 0.5±1.1 | <0.0001 | 11.9±26.7 | 4.3±14.2 | <0.0001 |
| COPD-OSA | 52 | 1.9±2.7 | 1.5±2.6 | 0.154 | 20.7±22.8 | 13.1±32.8 | 0.007 |
| NIV users | 259 | 1.2±1.8 | 0.9±1.9 | 0.0001 | 14.4±21.2 | 8.7±27.5 | <0.0001 |
| NIV non-users | 74 | 2.3±2.7 | 1.6±2.8 | 0.0056 | 26.9±41.5 | 13.0±24.3 | 0.0032 |

Values are mean ± standard deviation.

COPD, chronic obstructive pulmonary disease; NIV, non-invasive ventilation; ORRF, obesity-related respiratory failure; OSA, obstructive sleep apnoea.

**Table S2.** Survival outcomes in subgroups based on the indication for non-invasive ventilation (NIV), NIV use versus non-use at 12 months, and Scottish Index of Multiple Deprivation quintile.

|  | | **All** | **Diagnosis/indication for NIV** | | | **NIV at 12 months** | | **SIMD quintile** | |
| --- | --- | --- | --- | --- | --- | --- | --- | --- | --- |
|  | |  | **COPD** | **OSA/COPD** | **ORRF** | **Users** | **Non-users** | **1** | **2–5** |
| Descriptions | Participants, n | 362 | 89 | 56 | 217 | 274 | 88 | 163 | 197 |
|  | Median follow-up, days (IQR) | 1222  (869–1706) | 1071  (645–1402) | 1226  (873–1708) | 1328  (907–1799) | 1222  (869–2344) | 1232  (882–1709) | 1231  (874–1709) | 1219  (870–1699) |
| Time to readmission or death | Events, n | 218 | 78 | 41 | 99 | 147 | 71 | 94 | 123 |
|  | Median time to event, days (95% CI) | 946  (743, 1176) | 211 (140, 345) | 462  (306, 759 | 1655 (1363, N/A) | 1235 (932, 1629) | 374 (212, 684) | 932 (633, 1626) | 953 (686, 1212) |
|  | Hazard ratio (95% CI) | Ref. | 2.31 (1.78, 3.00) | 1.43 (1.02, 2.00) | 0.64 (0.50, 0.81) | Ref. | 2.11 (1.58, 2.8) | Ref. | 1.10 (0.84, 1.43) |
|  | Wald p-value |  | <0.0001 | 0.04 | 0.0002 |  | <0.0001 |  | 0.49 |
|  | Log rank p-value |  | <0.0001 | | | <0.0001 | | 0.5 | |
| Time to death | Events, n | 105 | 45 | 18 | 42 | 66 | 39 | 51 | 54 |
|  | Median time to event, days (95% CI) | NA | 1226 (1078, NA) | NA (1520, NA) | NA | NA | 1655  (1212, NA) | NA | NA |
|  | Hazard ratio (95% CI) | Ref. | 2.1 (1.48, 2.99) | 1.1 (0.67, 1.83) | 0.62 (0.44, 0.89) | Ref. | 2.25 (1.51, 3.34) | Ref. | 0.89 (0.60, 1.30) |
|  | Wald p-value |  | <0.0001 | 0.69 | 0.0096 |  | <0.0001 |  | 0.54 |
|  | Log rank p-value |  | <0.0001 | | | <0.0001 | | 0.5 | |
| Time to readmission | Events, n | 194 | 73 | 35 | 86 | 131 | 63 | 85 | 108 |
|  | Median time to event, days (95% CI) | 1176  (867, 1529) | 211 (140, 345) | 482 (374, 1456) | NA  (1546, NA) | 1529 (1176, NA) | 414 (227, 822) | 1304 (687, NA) | 1109 (822, 1458) |
|  | Hazard ratio (95% CI) | Ref. | 2.38 (1.82, 3.13) | 1.35 (0.94, 1.94) | 0.63  (0.49, 0.81) | Ref. | 2.06 (1.52, 2.78) | Ref. | 1.06 (0.80, 1.41) |
|  | Wald p-value |  | <0.0001 | 0.10 | 0.0003 |  | <0.0001 |  | 0.67 |
|  | Log rank p-value |  | <0.0001 | | | <0.0001 | | 0.7 | |

CI, confidence interval; COPD, chronic obstructive pulmonary disease; NA, not achieved; NIV, non-invasive ventilation; ORRF, obesity-related respiratory failure; OSA, obstructive sleep apnoea; Ref., reference; SIMD, Scottish Index of Multiple Deprivation.

**Table S3.** Survival outcomes in subgroups based on NIV usage at 12 months, and Scottish Index of Multiple Deprivation quintile.

|  | | Non-users | | Users | |
| --- | --- | --- | --- | --- | --- |
|  | | **SIMD 1** | **SIMD 2–5** | **SIMD 1** | **SIMD 2–5** |
| Descriptions | Participants, n | 40 | 47 | 123 | 150 |
|  | Median follow-up, days (IQR) | 1233 (876–1716) | 1226 (869–1701) | 1229 (874–1708) | 1223 (871–1700) |
| Time to readmission or death | Events, n | 32 | 38 | 62 | 85 |
|  | Median time to event, days (95% CI) | 206 (79, 1056) | 463 (255, 953) | 1388 (812, NA) | 1172 (788, 1599) |
|  | Hazard ratio (95% CI) | Ref. | 0.9 (0.56, 1.43) | Ref. | 1.2 (0.85, 1.64) |
|  | Wald p-value |  | 0.6 |  | 0.3 |
|  | Log rank p-value | 0.6 | | 0.3 | |
| Time to death | Events, n | 21 | 18 | 30 | 36 |
|  | Median time to event, days (95% CI) | 1335 (759, NA) | NA (1212, NA) | NA | NA |
|  | Hazard ratio (95% CI) | Ref. | 0.7 (0.35, 1.23) | Ref. | 1.1 (0.64, 1.68) |
|  | Wald p-value |  | 0.2 |  | 0.9 |
|  | Log rank p-value | 0.2 | | 0.9 | |
| Time to readmission | Events, n | 27 | 35 | 58 | 73 |
|  | Median time to event, days (95% CI) | 206 (79, NA) | 482 (262, 965) | 1840 (867, NA) | 1340 (1053, NA) |
|  | Hazard ratio (95% CI) | Ref. | 0.9 (0.57, 1.57) | Ref. | 1.1 (0.77, 1.54) |
|  | Wald p-value |  | 0.8 |  | 0.6 |
|  | Log rank p-value | 0.8 | | 0.6 | |

CI, confidence interval; COPD, chronic obstructive pulmonary disease; NA, not achieved; NIV, non-invasive ventilation; ORRF, obesity-related respiratory failure; OSA, obstructive sleep apnoea; Ref., reference; SIMD, Scottish Index of Multiple Deprivation
